# Automatic Identification of Upper Extremity Rehabilitation Exercise Type and Dose Using Body-Worn Sensors and Machine Learning: A Pilot Study

**DOI:** 10.1101/2020.04.17.20043869

**Authors:** Noah Balestra, Gaurav Sharma, Linda M. Riek, Ania Busza

## Abstract

Stroke is a major cause of adult-onset disability worldwide, and approximately 40% of stroke survivors have residual impairment in upper-extremity function. Prior studies suggest that increased participation in rehabilitation exercises improves motor function post-stroke,^1^ however further clarification of optimal exercise dose and timing has been limited by the technical challenge of quantifying exercise activities over multiple days. In this exploratory study, we assessed the feasibility of using body-worn sensors to track rehabilitation exercises in the inpatient setting, and investigated which recording parameters and data analysis strategies are sufficient for accurately identifying and counting exercise repetitions. MC10 BioStampRC® sensors were used to measure accelerometry and gyroscopy from arms of healthy controls (n=11) and patients with arm weakness due to recent stroke (n=13) while the subjects performed three pre-selected upper extremity exercises. Sensor data was then labeled by exercise type, and this labeled data set was used to train a machine learning classification algorithm for identifying exercise type. The machine-learning algorithm and a peak-finding algorithm was used to count exercise repetitions in non-labeled data sets. We achieved a repetition counting accuracy of 95.6 ± 2.4 % overall, and 95.0 ± 2.3 % in patients with arm weakness due to stroke. Accuracy was decreased when using fewer sensors or using accelerometry alone. Our exploratory study suggests that body-worn sensor systems are technically feasible, well-tolerated in subjects with recent stroke, and may ultimately be useful for developing a system to measure total exercise “dose” in post-stroke patients during clinical rehabilitation or clinical trials.

## Introduction

Each year, nearly 800,000 strokes occur in the US alone^2^, and approximately 40% of individuals with stroke are left with permanent functional disability^3^. In recent decades there has been major progress in acute stroke treatments and stroke survival^2^. In contrast, the field of neurorehabilitation has lagged behind, with only a limited number of interventions showing consistent effects across multiple trials and less than 10% of the American Heart Association (AHA) adult stroke rehabilitation guidelines based on strong (i.e. Class I or Level A) evidence.^3,4^ In recent years, there has been a call for improving the quality of rehabilitation research^4^ and introducing more qualitative outcome and motor function measures.

One aspect of rehabilitation and recovery which has previously eluded detailed quantification is the amount of repetitive exercises, or rehabilitation “dose”, that the patient engages in as part of their rehabilitation therapy. Classically, stroke rehabilitation researchers have used units of time spent in therapy when trying to account for dose of stroke rehabilitation exposure. However, studies measuring the actual amount of exercise performed during rehabilitation show that the same amount of time can represent a wide range of patient participation and number of repetitions1,^5^. This observed discrepancy demonstrates the potential risks of using “time spent in therapy” as a measure of rehabilitation dose. This variability in exercise dose measurement may contribute to unexplained variability in recovery outcomes between different patients. Furthermore, inaccurate rehabilitation exercise dose measurement makes it difficult to test hypotheses about the relationship between exercise type, dose, and functional recovery.

Prior studies examining the dose-response relationship in patients with stroke have suggested that a preferable measure of exercise dose would involve the counting of repetitions for each exercise performed in therapy^1^. However, manual repetition counting of patient exercises is laborious and error-prone. New technologies and sensors may enable automated systems for exercise repetition-counting. This could be accomplished by using optical motion capture technology to track patient arm movements. However, this method has classically required multiple camera angles and rigid body surface markers, and is therefore predominantly used for kinematic assessment rather than exercise quantification^6^. While newer artificial intelligence – based motion analysis software systems no longer require markers to identify limb movements^7^, they would still require constant video monitoring and proper visibility to track a patient’s exercises throughout the day.

In recent years, new sensor technologies and more accessible machine learning programs has led to a surge of interest in automatic sensor-based systems for movement classification. Body-worn sensors have been used to identify specific movement patterns of the wearer in both healthy controls and patients with illness, including patients with neurological diseases^11^.

We therefore conducted a pilot-study in the inpatient setting to assess the ability to automatically measure exercise repetition “dose” using body-worn sensors. Specifically, we recruited healthy controls and patients with hemiparesis due to stroke admitted to our hospital’s stroke inpatient and acute rehabilitation units for our pilot-study. Over the course of several hours, participants wore superficial sensors (BioStampRC, MC 10 Inc., Lexington, MA, USA) and performed several sets of 3 pre-defined upper extremity exercises. Using accelerometry and gyroscope data collected from the sensors, we compared the effect of sensor placement, sensor data, and data analysis strategies on our system’s ability to (1) automatically categorize exercise type and (2) accurately count number of repetitions of the specific exercise type, in new (unlabeled) data sets.

## Methods

### Participants and Inclusion/Exclusion Criteria

Subjects were recruited through posted flyers and emails and from inpatient rehabilitation, stroke recovery, and neurosurgery units at the Strong Memorial Hospital in Rochester, NY. The study protocol was approved by the Institutional Review Board of the University of Rochester and all subjects signed an informed consent document prior to starting study procedures. Inclusion Criteria (for subjects with history of stroke) included having moderate arm weakness (Medical Research Council (MRC) strength scale score: 3-4) due to recent stroke. Patients with both ischemic and hemorrhagic strokes were recruited for this study. Exclusion criteria (for both healthy controls and subjects with recent stroke) included chronic arm injury or pain. Patients with history of recent stroke were also excluded if they had severe arm weakness (MRC≤2), aphasia/cognitive impairment such that the patient is not independently making choices about their healthcare decisions, or if their health care provider determined they were unable to safely participate in rehabilitation therapies.

### Sensor Placement and Data Acquisition

Three MC10 BioStampRC® wearable sensors were placed on the subject’s affected arms to record a combination of either triaxial accelerometry and electromyography or triaxial accelerometry and gyroscope data during three prompted upper-extremity exercises and resting periods. In this manuscript, only results using accelerometry and gyroscopy data are discussed.

One sensor each was placed on the upper arm (volar surface of the brachium), forearm (medial volar surface), and hand (dorsal surface) (Fig 1). The forearm sensor was originally placed on the medial forearm over the bulk of the wrist flexor muscle group (Fig 1A). However, when put into practice with inpatient subjects, this sensor placement was periodically inconvenient due to the presence of peripheral intravenous lines or large, painful bruises that formed after the removal of such lines from the antecubital region. Thus we also collected data a more lateral position on forearm, over the wrist extensor muscle group (Fig 1B), to ensure that the system could still be applied for a patient if the antecubital fossa region were unavailable for sensor placement (for example, when a line is placed in the paretic arm in the setting of emergency medical treatment or because both arms are paretic).

**Figure 1:**
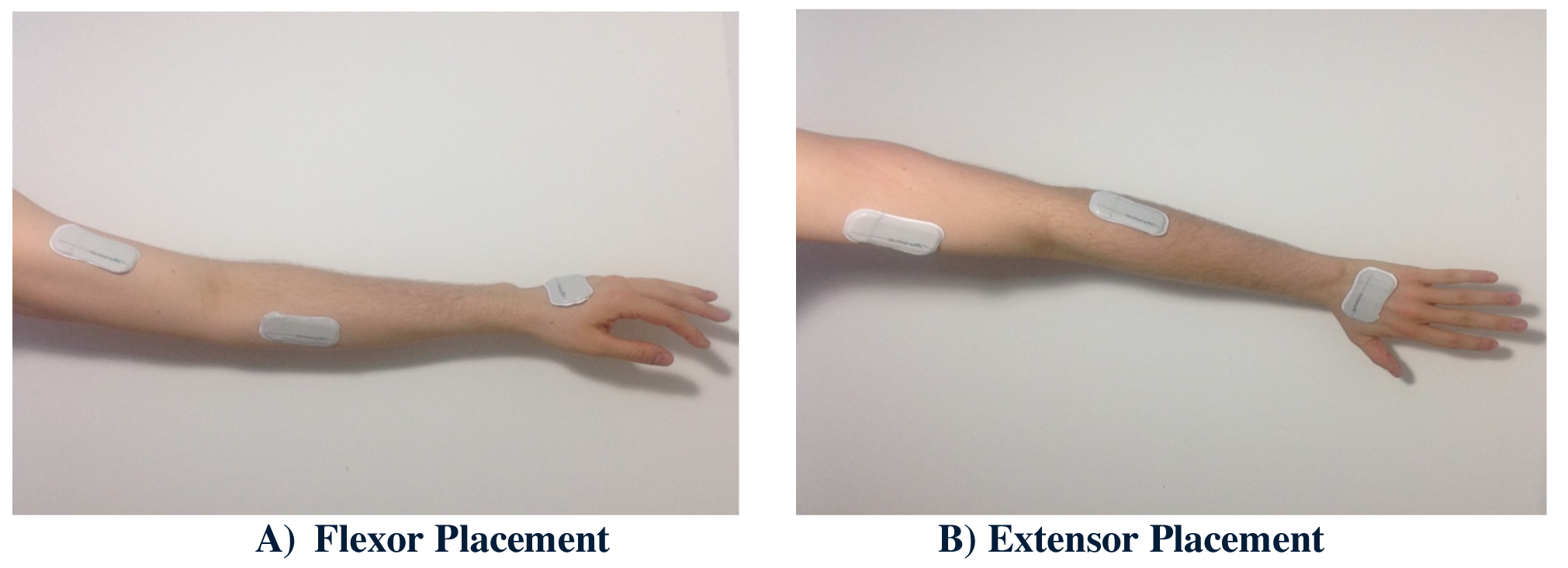
Illustration of sensor placement on the left arm. Sensors were placed on the upper arm, dorsal surface of the hand, and over either (A) the wrist flexor muscle group or (B) the wrist extensor muscle group of the forearm. See text for more details.

### Exercise Protocol

The subjects were received verbal instructions and visual demonstrations on how to perform three exercises (Fig 2.):

1. Exercise 1: flexion/extension of the elbow
2. Exercise 2: supination/pronation of the forearm
3. Exercise 3: extension/flexion of the wrist

**Figure 2:**
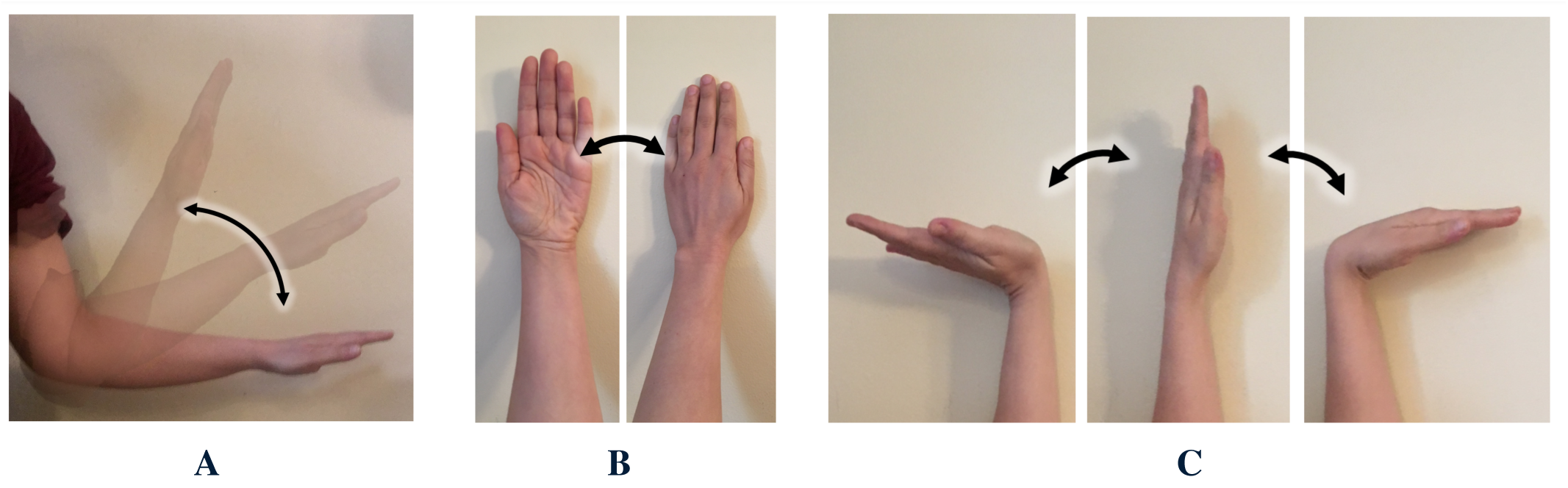
Illustration of upper-limb exercises. (A) Exercise 1 (elbow flexion and extension), (B) Exercise 2 (forearm supination and pronation), and (C) Exercise 3 (wrist extension and flexion).

Patients were asked to perform 20 repetitions of each exercise per session, with a total of three exercise sessions separated by at least 20 minutes. To ensure accurate movements, study personnel supervised the exercise activities during the exercise session and provided additional demonstrations of the correct exercise movements as needed. They also tagged the start and stop of each exercise type on the MC10 application, so that the “true” activity was labeled at the correct time on the data recording. Sensors were kept on subjects’ arms for one to three hours in order to record data corresponding to resting and non-exercise movement patterns and confirm that the algorithm can effectively discriminate and ignore non-exercise arm movements.

Patients were allowed to perform any paretic arm movement they wished during the resting periods, but were asked to avoid movements that replicated any of the three study exercises. Recording sessions were timed such that the subject did not have occupational or physical therapy rehabilitation sessions during the data recording period.

### Data Processing

Data were recorded simultaneously from the three sensors. Since data sets from different sensors are not perfectly synchronized and are sampled at different sampling frequencies, post-processing was required to ensure each raw data set could be used to train the machine learning-based classification algorithm. MATLAB and Delimit software programs were used for all data processing. The first step involved linear interpolation of the data to match sensor sampling frequencies, which may differ depending on the recording mode. This was done in MATLAB using the interp1 function. In the second step, data was synchronized by the first Unix timestamp value that was associated with each period in the exercise session. The data was then labeled according to Unix timestamp intervals which were created by a start/stop timer included in the MC10 application. Data sets were filtered using a bandpass filter (0.1 to 1.5 Hz) to remove high-frequency noise and wandering baselines due to gravity signatures (changes in baselines due to the force of gravity acting on the sensors differently, depending on their orientation) then z-score normalized in MATLAB using the normalize function.

### Classification, Data Extraction, and Repetition Enumeration Using Peak Finding

Once all data sets were synchronized, labelled, filtered, and normalized, the Fine *k* nearest neighbor (*k-*NN) classification algorithm provided by MATLAB was used to classify the data using a leave-one-out cross validation method. This was accomplished by training the algorithm on all datasets except for one, and then testing the algorithm on the “left-out” dataset. This test produced a set of algorithm-predicted labels for each row of the “left out” dataset. The Fine *k-*NN algorithm was utilized over other possible classification algorithms because when applied to the data in MATLAB’s classification learner add-on, the Fine *k-*NN algorithm achieved the highest reported classification accuracy (Table 1).

**Table 1:** Mean classification accuracy rates using different Classification Algorithms

| Algorithm | True Positive Rates<br>(%, Averaged Across Classes) | False Discovery Rate<br>(%, Averaged Across Classes) |
| --- | --- | --- |
| **Fine KNN** | **98.5** | **0.7** |
| Medium KNN | 94.1 | 2.7 |
| Coarse KNN | 84.5 | 5.0 |
| Cosine KNN | 92.5 | 7.5 |
| Cubic KNN | 94.1 | 3.1 |
| Weighted KNN | 95.7 | 1.0 |
| Subspace KNN | 92.9 | 0.5 |
| Fine Tree | 79.2 | 11.8 |
| Linear SVM | 25.0 | 1.9 |
| Quadratic SVM | 82.9 | 5.4 |
| Cubic SVM | 87.9 | 3.9 |
| Fine Gaussian SVM | 74.5 | 2.3 |

To calculate the number of repetitions a subject performed of each exercise, data corresponding to Exercise 1, Exercise 2, and Exercise 3 were separately extracted from each dataset according to the column of algorithm predicted discrete labels, and new tables of each activity were created for each subject. The MATLAB findpeaks function was then applied to different variable signals in each of these extracted activity tables to count the number of repetitions for each activity. For the Exercise 1 (elbow flexion/extension) tables, the x-axis accelerometer data from the forearm sensor was used, because clear peaks representing Exercise 1 repetitions are found in this data. For the same reason, the y-axis and x-axis gyroscope signal from the dorsal hand sensor were used to count repetitions of Exercise 2 and 3, respectively. An example of this process, as well as the previously mentioned data pre-processing methods, are shown in Figure 3.

**Figure 3:**
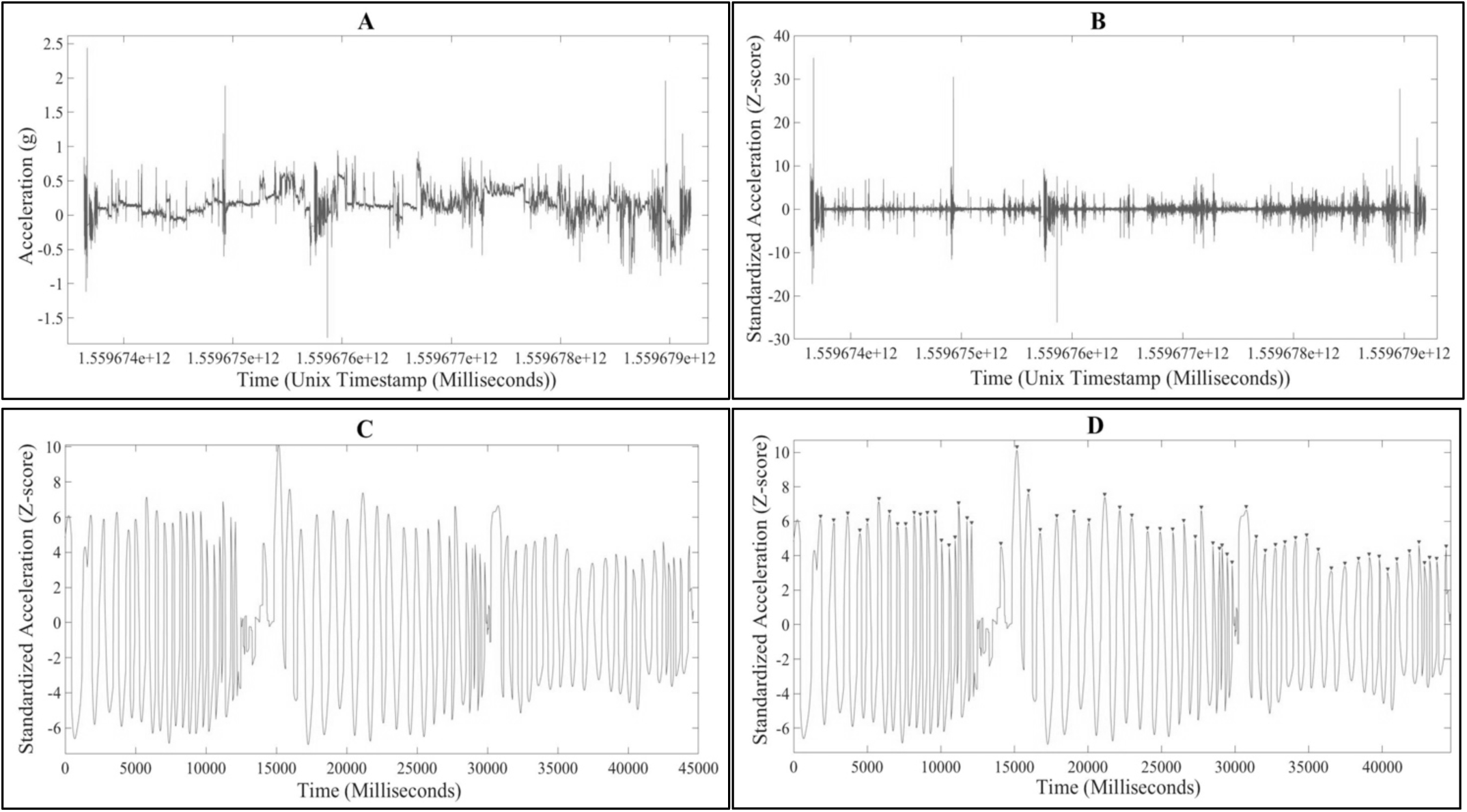
Outputs of data pre-processing, automatic labelling, and peak finding functions. A) Raw x-axis accelerometer data from the forearm sensor, from subject #6’s dataset. B) Data (from A) after high pass filtering and normalization. C) Data classified as “Exercise 1” and extracted by the algorithm from the normalized, filtered data. D) The output of the findpeaks function when used on the extracted Exercise 1 data. Each triangular marker represents a found peak, which the findpeaks function also counts and reports.

### Peak Counting Accuracy Calculation

To determine peak counting accuracy, the system’s estimate of the number of repetitions performed by the patient for each exercise, *N*_automatic_, was compared to the repetition number as reported by the study personnel, *N*_manual_. To calculate this accuracy, the percent error was calculated for each count, and subtracted from 100% according to the following formula:

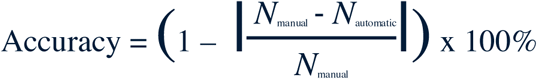

## Results

### Participants

11 of the subjects enrolled in this study were healthy controls with no arm weakness (average age: 43 years old, range: 20 to 79 years old). 23 subjects with recent stroke initially consented to the study, but then 4 were excluded because they were unable to perform the three selected exercises (either due to somnolence or due to the affected arm being too weak to engage in multiple repetitions of the preselected exercises). Three subjects had only one or two sensors on throughout the recording period due to limitations in numbers of sensors, and an additional three subjects had a sensor fall off during the session. In the end, 13 subjects with recent stroke completed the study. Of the 13 subjects whose data were used, mean age was 70 years old (range 40 to 90 years old), mean MRC scale score was 3.83, and mean time since stroke onset was 7.8 days.

### Study Protocol Deviations, Technical Issues, and Adverse Events

Study sessions for monitored for complications such as accidental sensor removal during recording periods, skin irritation due to prolonged/repeated sensor placement, and allergic reactions to the adhesive material on the sensors. All subjects completed their testing time, no subjects reported skin irritation or allergic reactions to the sensors at any point during their participation in the study, and subjects typically found the sensors to be unnoticeable in terms of their comfort. However, there were six recording sessions in which accidental sensor removal did occur due to adhesive failure. When accidental sensor removal did occur, the resultant data set was either not used or re-recorded from the subject at a different time.

### Comparison of Classification Accuracy between MATLAB Classification Algorithms

To determine which MATLAB classification algorithm would be best for the automatic repetition counting system, each possible MATLAB classification algorithm was applied to each of the data sets that included gyroscope and accelerometer data, and the average classification accuracies were compared, shown in Table 1 below. The Fine KNN or *k*-NN algorithm achieved the highest classification accuracy rates out of the relevant options.

### Comparison of Repetition Counting Accuracy for Control and Stroke Patient Data Sets

Repetition Count Accuracy was compared between data obtained from healthy controls and data from stroke patients, who may often have higher variability in exercise performance due to weakness. With only accelerometer data used, the mean accuracy for healthy control data sets is **76.1** ± **21.5 %**, while the mean accuracy for the stroke patient data sets is **72.3** ± **24.8 %**. There was no significant difference between the two groups (*p* = .67). For the data sets with gyroscope data, the mean accuracy for healthy control data sets is **96.2** ± **2.4 %**, and the mean accuracy for the stroke patient data sets is **95.0** ± **2.3 %**. Again, no significant difference was found between the control and stroke patient groups *(p =* .48).

### Comparison of Data Acquisition and Data Analysis Strategies

To determine if recording gyroscope data in addition to accelerometer data improved accuracy, the accuracy of data sets with gyroscope data was compared to those without. The mean accuracy for subject datasets with only accelerometer data is **74.6** ± **22.8 %**, while the mean accuracy for datasets that had both accelerometer and gyroscope data is **95.6 ± 2.4 %**.

To ensure that this large discrepancy in accuracy is due to the effect of adding gyroscope data to the analysis, and not due to variability between subjects, accuracy with and without gyroscope data were compared for each subject that had both accelerometer and gyroscope data collected. The overall mean accuracy without gyroscope data for these subjects is **84.3** ± **18.0 %**, compared to the mean accuracy for these subjects with gyroscope data, **95.6 ± 2.3 %**. In every subject that had both accelerometry and gyroscopy data collected during their study session, adding gyroscope data to accelerometry data improved repetition counting accuracy (Fig 4).

**Figure 4:**
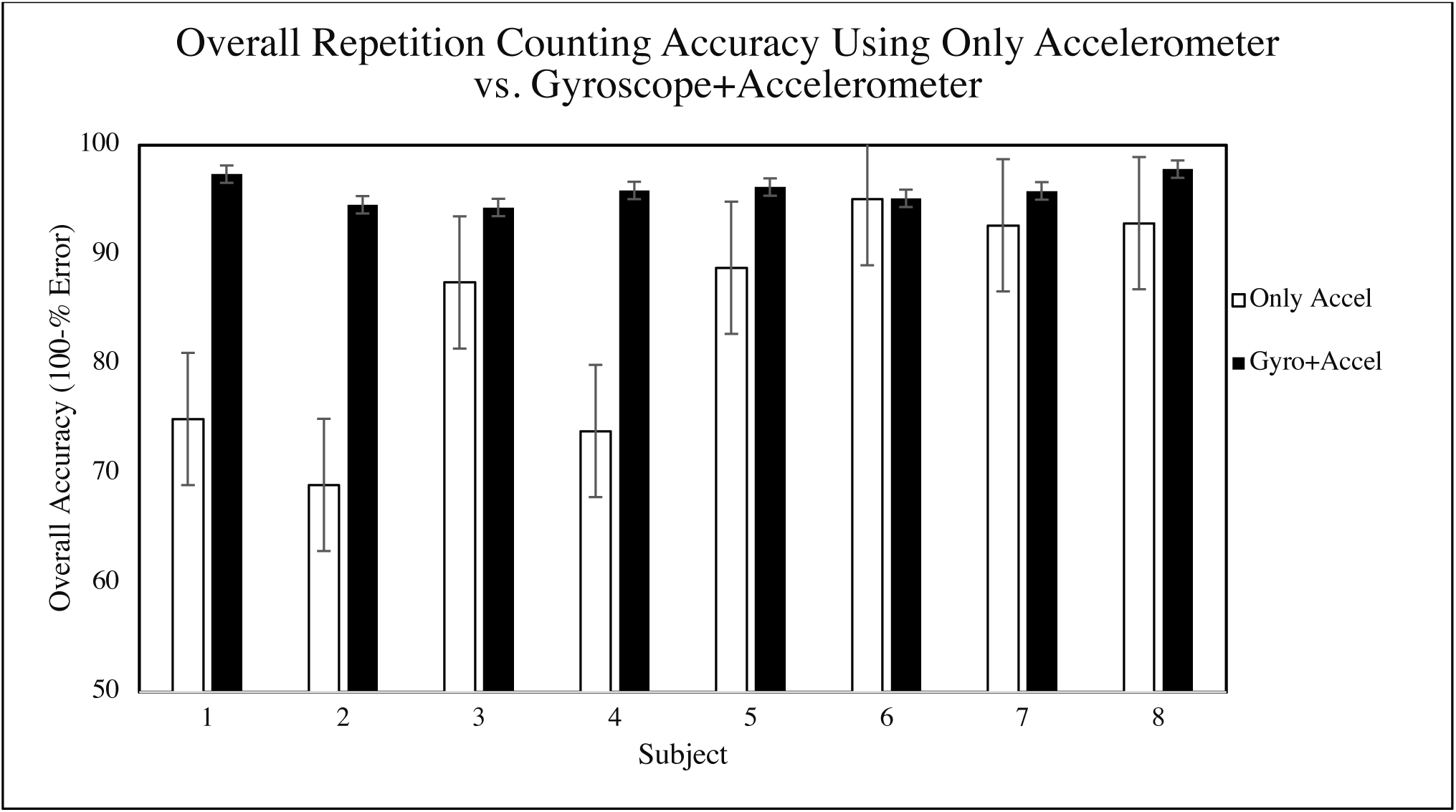
Peak counting for every subject from which gyroscope data was recorded, with and without gyroscope data implemented. Error bars represent the 95% confidence interval for each overall accuracy calculation. Subjects 1-4 were patients with arm weakness due to recent stroke.

Even in relatively weak patients (i.e. MRC strength scale of 3) we found repetition accuracy of >94% when gyroscope data from the dorsal hand was added (Table 2)

**Table 2:**
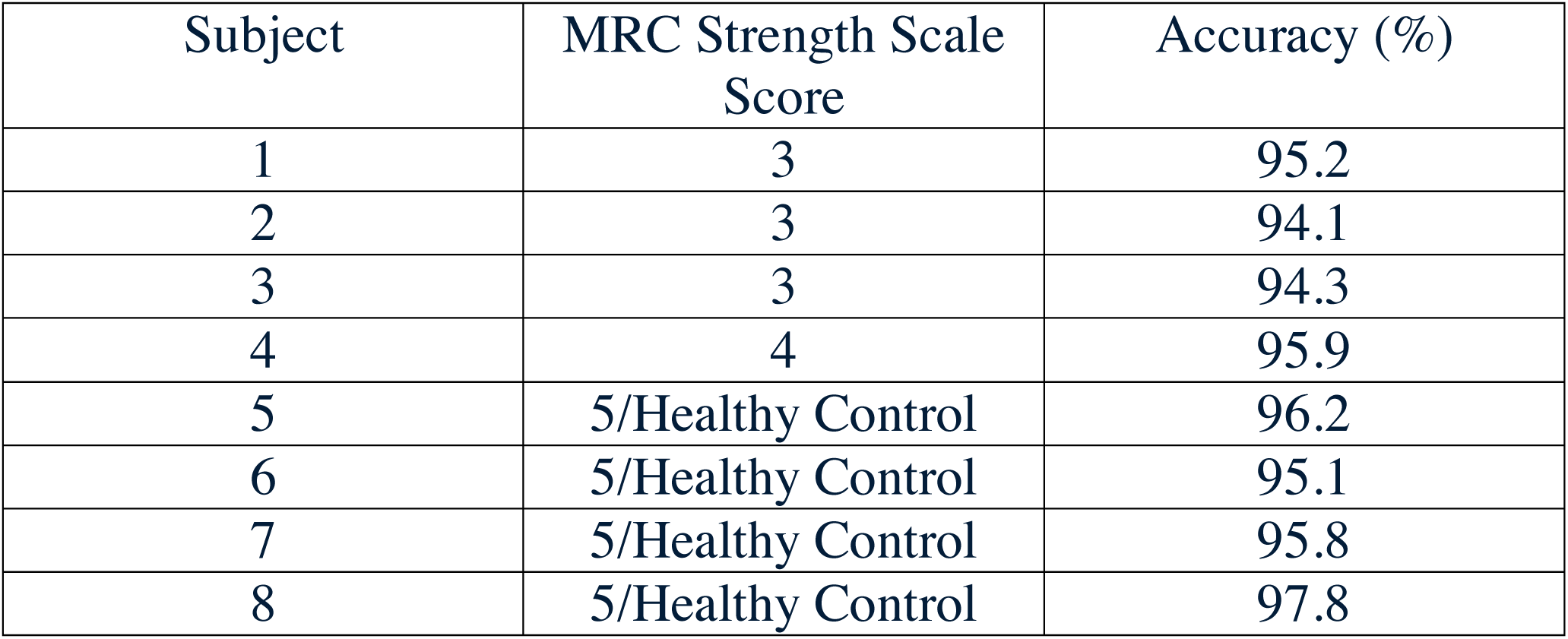
Overall accuracy (averaged from each exercise) for each subject from which gyroscope data was recorded. The Medical Research Council (MRC) scale is a scale used to determine relative strength. A score of 3 indicates an ability to move the affected arm against gravity but not resistance, a score of 4 indicates an ability to move against gravity and some resistance, and a score of 5 indicates normal arm strength.

### Effect of Number of Sensors

Using the parallel coordinates plot in MATLAB, we evaluated the relevance of the different variables in terms of classification accuracy (discrimination between the exercises) and found that the gyroscope + accelerometer data from the hand sensor and the accelerometer data from the forearm sensor are the most important variables for activity classification. Additionally, the findpeaks function performs most accurately when used on the x-axis accelerometer data from the forearm sensor or x-axis gyroscope data from the hand sensor for Exercise 1, the y-axis gyroscope data from the hand sensor for Exercise 2, and the x-axis gyroscope data from the hand sensor for Exercise 3. These findings suggest a relative sensor importance of Dorsal Hand > Forearm > Upper Arm (Brachium) sensor. Accordingly, when reducing the number of sensors, we decided to remove the upper arm sensor when limiting to two sensors and the upper arm and forearm sensor when limiting to one sensor. The additional testing to determine the accuracy of the systems with reduced sensor data was only performed for subjects from which gyroscope data was recorded. The mean accuracy for the system is **95.6 ± 2.4 %** when incorporating data from 3 sensors, **92.7** ± **6.6 %** when using 2 sensors, and **85.1** ± **11.9** % when using 1 sensor. (Fig 5)

**Figure 5:**
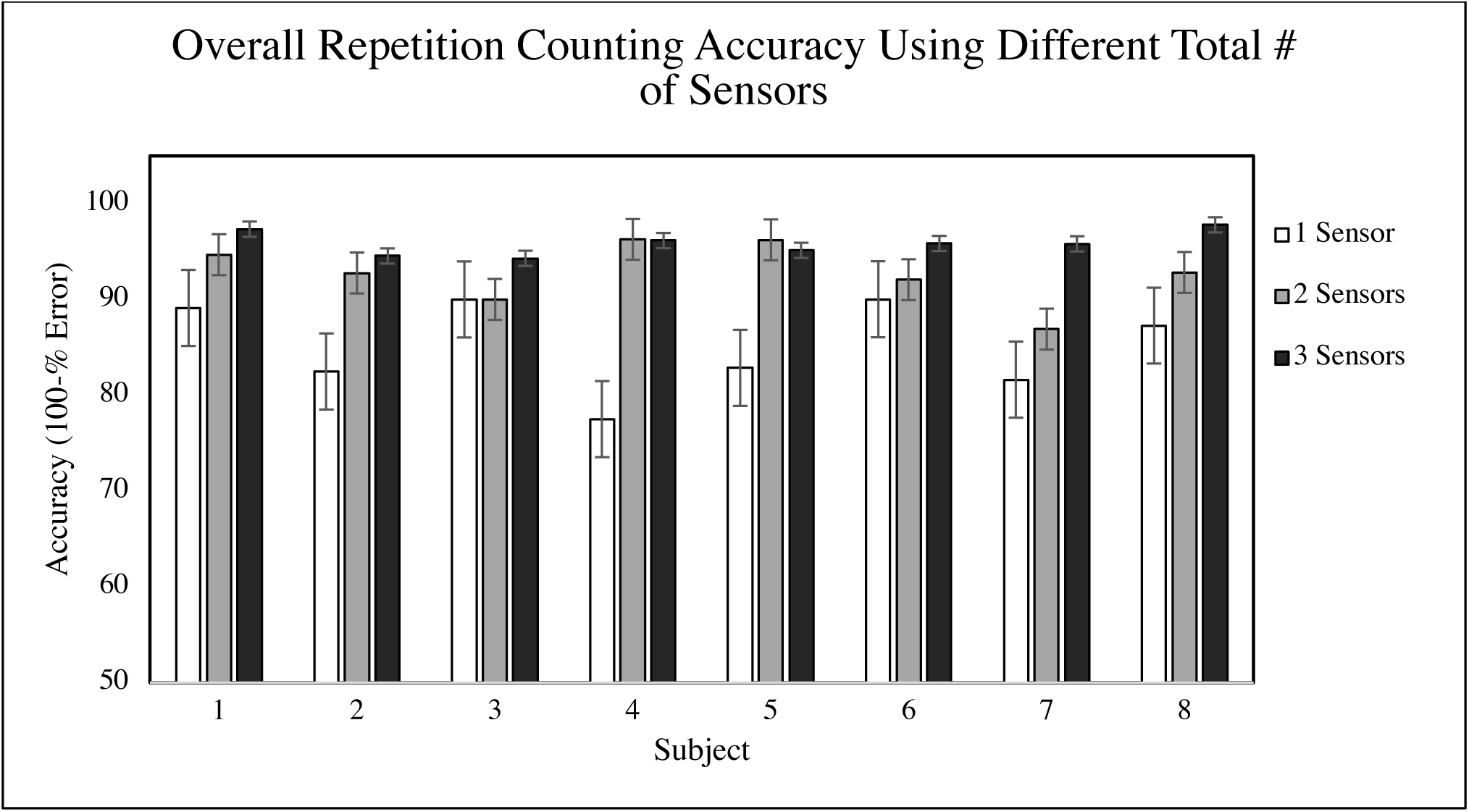
The peak counting accuracy for each subject from which gyroscope data was recorded, varying with number of sensors used. Subjects 1-4 were patients with arm weakness due to recent stroke, subjects 5-8 were healthy controls. Error bars represent the 95% confidence interval for each overall accuracy calculation.

### Effect of Forearm Sensor Placement

To ensure that the forearm sensor could be placed on either the flexor or extensor digitorum muscles and achieve similar levels of accuracy, the overall accuracies for all subjects with flexor and extensor sensor placements were compared. Since a larger number of the datasets that include gyroscope data used the flexor digitorum sensor position, only the accelerometry data from all datasets was used. The mean accuracy across all subjects with the forearm sensor in the flexor position is **76.8** ± **19.1 %**, and from the extensor position, **72.0** ± **26.6 %**. In order to determine if this difference in accuracy is statistically significant, a 2 sample T-test was performed (*p* = .58). This result indicates that there is not a statistically significant difference between the two sensor placements, and either forearm sensor position is viable for the system.

## Discussion

The goal of this study was to assess the feasibility of using body-worn sensors to automatically count exercise repetitions in the inpatient stroke and acute rehabilitation setting, and to investigate the effect of using different recording parameters on repetition count accuracy.

In our 13 healthy controls and 13 inpatient hospital subjects with recent stroke, the sensors were well tolerated and no subjects reported skin irritation or other side effects. However, there were multiple instances of skin adherence failures, when the adhesive sticker failed to keep the sensor on the subject’s arm for the entire duration of the study. Possible future solutions to this issue includes using more adherent adhesives (although this will likely also increase patient discomfort with sensor removal). We have also found self-adherent (cohesive) bandages helpful when used in addition to the sensor adhesives.

Reducing the number of sensors per arm from 3 to 2 or 1 significantly reduced repetition counting accuracy, while adding gyroscope data collection significantly improved accuracy – especially in subjects with arm weakness due to stroke. In these patients, adding gyroscope recording to the dorsal hand sensor improved repetition count accuracy by over **10% (84.3 to 95.6%)**. Healthy subjects did not show such a large improvement in accuracy with the addition of gyroscope data which may due to a ceiling effect as healthy subjects had higher repetition accuracy with accelerometry alone. This may be because gyroscope data measures rotation about an axis, and like many exercises practiced by patients with stroke as part of their rehabilitation regimen, the 3 exercises we chose for our study are rotational “range of motion” exercises. This suggests that gyroscopy is likely an important measurement to capture in future studies monitoring stroke rehabilitation efforts. Unfortunately, adding gyroscope data in addition to accelerometer data reduces the recording time of the current sensors from 21 hours to only 3 hours due to battery limitations (if both data are recorded with a 62.5 Hz sampling rate). This means that with gyroscope data being recorded, the sensors would have to be replaced and recharged once every three hours for each patient, which would make longer-term monitoring efforts infeasible until better sensor battery conserving solutions are found.

On all subjects from whom gyroscope data was recorded, an overall repetition-counting accuracy of at least 94.1% was achieved, even for patients with an MRC strength scale score of 3/5. The system’s repetition counting was more accurate when gyroscope data was recorded and implemented in addition to accelerometer data. The system’s repetition counting became less accurate as fewer sensors’ inputs were utilized, and the best performance was achieved using all three sensors’ data.

While the results of this study are promising, there are several limitations. First, the system has only been tested on three basic exercises in terms of classification and repetition counting ability. Secondly, all incorporated stroke patient data was collected from subjects with an MRC strength scale of 3 or greater (moderate arm weakness or better), which does not represent the entire population of patients with stroke. Therefore, the system’s ability to successfully quantify exercise dosage of a wider range of upper extremity exercises and/or from patients with greater arm weakness is uncertain. The method for counting exercise repetitions was developed using the readily available Fine *k* nearest neighbor algorithm and could be refined by using alternative time-series analysis approaches that can take into account the temporal progression of movements involved in an exercise.

In recent years, multiple studies have been published reporting the use of similar sensor systems for automated movement tracking, with the majority of these studies focusing on the classification of whole-body movements, such as standing, sitting, walking, ascending/descending stairs, playing sports, cycling, and others.^9,10^ Classification of upper extremity movements have been more limited in scope and/or application of the system. Biswas et al. and Bochniewicz et al. built classification algorithms for arm exercises and applied them to stroke patient data, but did not attempt to create a repetition quantifying system to measure exercise dose.^11,12^ Other examples, such as Zhang et al. and Crema et al. created arm exercise classification and repetition counting systems, but did not apply them to data from patients.^13,14^ The most recent study on this topic, Guerra et. al, developed a movement classification system that could be used to enumerate repetitions and applied the system to stroke patient data, but focused on the classification of movement primitives (components of arm movements that cannot be broken down further) rather than the exercises themselves.^15^ To our knowledge, this study is the first to explore the application of upper extremity exercise classification and repetition counting on exercise data in inpatients with recent stroke.

In summary, this work suggests that using wearable sensors in the inpatient stroke and acute rehabilitation setting is feasible and has the potential of creating an automated system to quantify individual rehabilitation therapy dose. Future work is needed to expand the range of rehabilitation activities identified by this system, and improve sensor adherence and battery life. Ultimately such a system may contribute to answering key questions about how patient exercise “dose” in the acute/subacute period post-stroke affects final motor outcomes, provide a system for providing patient feedback on how their efforts compare to target doses, and improve patient post-stroke function.

## Data Availability

Sensor data for this paper will be made available via request to the authors

## Acknowledgments

We would like to thank Simon Carsen, Tanzeem Choudhury and Kyle Choi for their contribution of advice and support, and the URMC neurology department residents, nursing staff, and occupational and physical therapists for their input, assistance with recruiting subjects, and their accommodation of the collection of data for the study.

